# Interplay between Tumor Mutational Burden and Mutational Profile and its effect on overall survival: A Post Hoc Analysis of Metastatic Patients Treated with Immune Checkpoint Inhibitors

**DOI:** 10.1101/2022.04.10.22273664

**Authors:** Camila B. Xavier, Carlos Diego H. Lopes, Beatriz M. Awni, Eduardo F. Campos, João Pedro B. Alves, Anamaria A. Camargo, Gabriela D. A. Guardia, Pedro A. F. Galante, Denis L. Jardim

## Abstract

**Purpose:** Solid tumors harboring tumor mutational burden (TMB) ≥ 10 mutations per megabase (mut/Mb) received agnostic approval for pembrolizumab. However, TMB cut-off alone is not a predictor of overall survival (OS). This work aims to analyze the somatic mutational profile’s influence in the outcomes of patients with TMB-high tumors treated with immune checkpoint inhibitors (ICIs).

**Methods:** This post-hoc analysis evaluated clinical and molecular features of 1,661 patients with solid tumors treated with ICIs. We performed OS analysis for TMB thresholds of ≥ 10, ≥ 20, and < 10 mut/Mb. For a TMB ≥ 10mut/Mb cutoff, we assessed OS according to mutational profile. For genes exhibiting a correlation with OS (P < 0.05) at the univariate assessment, we conducted a Cox multivariate analysis adjusted by median TMB, sex, age, microsatellite instability (MSI), and histology.

**Results:** 1,661 patients were investigated, and 488 harbored a TMB ≥ 10 mut/Mb (29.4%). The median OS was 42 months for TMB ≥ 10 or 20 mut/Mb, and 15 months for TMB < 10 mut/Mb (P < 0.005). In patients harboring TMB ≥ 10mut/Mb, mutations in E2F3 or STK11 were correlated with worse OS, and mutations in *NTRK3, PTPRD, RNF43, TENT5C, TET1* or *ZFHX3* with better OS. These associations were confirmed by univariate and multivariate analyses (P < 0.05). Melanoma histology and TMB above the median endowed patients with better OS (P < 0.05). MSI status, age, and gender did not have a consistent statistically significant effect on OS

**Conclusion:** Combining TMB information and mutation profiles in key cancer genes can be used to better qualify patients for ICI treatment and predict their OS.

**CONTEXT SUMMARY:** *Key objective:* Tumor mutational burden (TMB) of ≥ 10 mutations per megabase (mut/Mb) grant agnostic indication of pembrolizumab for advanced solid tumors’ treatment, however a substantial number of patients do not respond to therapy. This work aims to analyze the somatic mutational profile’s influence in the outcomes of patients with TMB ≥ 10mut/Mb tumors treated with ICIs.

*Knowledge generated:* Mutation profile can modify survival outcomes to ICIs in patients with TMB ≥ 10 mut/Mb. Mutations in *E2F3* or *STK11* correlate with worse OS, while mutations in *NTRK3, PTPRD, RNF43, TENT5C, TET1* or *ZFHX3* correlate with better OS in TMB-high patients receiving ICIs.

*Relevance:* We found that the combination of a high TMB and the somatic mutational profile in key cancer genes can be decisive in better qualify patients for ICI treatment.

## INTRODUCTION

Tumor mutational burden (TMB) has been correlated with the response to immune checkpoint inhibitors (ICIs) in a retrospective cohort including 1,661 patients treated with ICIs. Among all patients, higher somatic TMB, defined as the highest (20%) in histology, was a predictor of better overall survival (OS)^1^. Subsequently, a prospective analysis from the phase II KEYNOTE-158 trial stated that a TMB of at least 10 somatic tumor mutations per megabase (mut/Mb) was associated with a higher proportion of objective response rates (ORR) to pembrolizumab monotherapy^2^. These results led to the FDA agnostic approval of pembrolizumab for TMB-high (≥ 10mut/Mb) patients. However, 42% of patients presenting with high TMB do not respond to ICIs, indicating the need for better patient selection in this setting. Currently, a variety of clinical and molecular factors may have important roles in modulating the tumor response to ICIs^3^. Factors such as MHC diversity^4,5^ and concomitant molecular alterations influence TMB as a biomarker for immunotherapy. To better qualify patients with TMB ≥ 10 mut/Mb for ICI treatment, we investigated their mutation profiles and correlated molecular alterations with their survival outcomes when treated with ICIs.

## METHODS

### Study Design

We collected genomic and survival data from 1,661 patients^1^ and retrieved their mutation profiles (MSK-IMPACT). All the data collected was available on the public database https://www.cbioportal.org/^6,7^ MSK-IMPACT assay identifies somatic exonic mutations in a predefined subset of 468 cancer-related genes (earlier versions included 341 or 410 genes), by using both tumor-derived and matched germline normal DNA. TMB was determined by the number of nonsynonymous somatic mutations. Firstly, a validation analysis correlated predefined percentiles used in the original publication^1^ with the absolute thresholds of ≥ 10, ≥ 20, and < 10 mut/Mb. Further, for a TMB ≥ 10mut/Mb cutoff, we selected mutations that occurred in at least 5 patients and assessed OS according to somatic mutational profile in key cancer genes. For genes mutations exhibiting a positive correlation with OS (P < 0.05) at the univariate assessment, we conducted a Cox multivariate analysis adjusted by median TMB, sex, median age, microsatellite instability (MSI) status, and histology. Since MSI status was not available in the current database, an individual assessment of somatic mutations in *MLH1, MSH2, MSH6, PMS2*, and *SETD2* was used as a surrogate for MSI estimation^8^. A flowchart encompassing the study design is available in **Appendix Fig A1**.

### Statistical Analysis

Overall survival (OS) for all patients who received at least one dose of ICIs was estimated using the Kaplan-Meier method. The Cox regression model was used to define the hazard ratios (HRs) for death, and a log-rank test was used to compare the results (95% confidence intervals for all analyses). The Python Lifelines package (version 0.26.4)^9^ was used for Kaplan– Meier and Cox analyses.

For a TMB ≥ 10mut/Mb cutoff (N = 488), we assessed OS regarding the mutational status of each gene mutation found in at least 5 patients (N = 392). For all genes exhibiting a correlation with survival considering a standard alpha-error level (P < 0.05), a Cox multivariate analysis was also conducted using Reboot^10^. The adjustment variables included sex, median age, microsatellite instability (MSI) status, TMB under or above the median TMB of the cohort (20 mut/Mb), and histology (non-small cell lung cancer (NSCLC), melanoma, bladder cancer, and colorectal cancer).

## RESULTS

### Patients’ characteristics

A total of 1,661 patients (11 cancer types) were included. The median follow-up was 19 months (range 0–80). The top three more incident tumors were NSCLC (21.1%), melanoma (19.3%), and bladder cancer (12.9%). Of all samples, 488 (29.4%) harbored a TMB of ≥ 10 mut/Mb. When the original criteria of top 20% in each histology was adopted, only 332 (20%) patients were considered TMB-high. Of these patients, 279 (84%) presented TMB ≥ 10 mut/Mb. MSI surrogates were detected in 42 (29.1%) of the cases. The ICI class received was anti-PD-(L)1 in 78.7%, anti-CTLA-4 in 6%, and ICI combination in 15.4% of the cases. Genes with most frequently harboring genomic alterations (incidence >10% of all samples) were *TP53, TERT, KMT2D, KRAS, PIK3CA, ARID1A, NF1*, and *PTPRT*. Sample characteristics are summarized in **Table 1**. After a maximum follow-up of 80 months, the median OS was 42 months for both TMB ≥ 10mut/Mb and TMB ≥ 20 mut/Mb, and 15 months for TMB < 10 mut/Mb, multivariate log-rank P < 0.005. The HRs for death were HR 0.44 (95% CI 0.34 - 0.56) and 0.57 (95% CI 0.49 - 0.67) for TMB ≥ 20 mut/Mb and ≥ 10 mut/Mb, respectively (P < 0.005). No difference was observed in death risk between cohorts with TMB 10 mut/Mb or less and 1 mut/Mb or less (HR 0.96; 95% CI 0.74 – 1.24; P = 0.73). **Figure 1**.

**Table 1.** Sample characteristics.

|  | **TMB < 10 mut/Mb** | **TMB ≥ 10 mut/Mb** | **Total** |
| --- | --- | --- | --- |
| **N (%)** | 1,173 (70.6) | 488 (29.4) | 1,661 (100) |
| **Tumor Type N (%)** |  |  |  |
| **NSCLC** | 235 (20) | 115 (23.6) | 350 (21.1) |
| **Melanoma** | 168 (14.3) | 152 (31.1) | 320 (19.3) |
| **Bladder** | 126 (10.7) | 89 (18.2) | 215 (12.9) |
| **Renal cell carcinoma** | 149 (12.7) | 2 (0.4) | 151 (9.1) |
| **Head and neck cancer** | 111 (9.5) | 28 (5.7) | 139 (8.4) |
| **Esophagogastric cancer** | 107 (9.1) | 19 (3.9) | 126 (7.6) |
| **Glioma** | 108 (9.2) | 9 (1.8) | 117 (7) |
| **Colorectal cancer** | 64 (5.5) | 46 (9.4) | 110 (6.6) |
| **Unknown primary cancer** | 64 (5.5) | 24 (4.9) | 88 (5.3) |
| **Breast cancer** | 41 (3.5) | 3 (0.6) | 44 (2.6) |
| **Skin (non-melanoma)** | 0 (0) | 1 (0.2) | 1 (0.1) |
| **Median age (years)** | 62 | 66 |  |
| **Sex** |  |  |  |
| **Female** | 447 (38.1) | 180 (36.9) | 627 (37.8) |
| **Male** | 726 (61.9) | 308 (63.1) | 1034 (62.2) |
| **MSI (%)** | 72 (6.1) | 142 (29.1) | 214 (12.9) |
| **ICI type** |  |  |  |
| **ANTI-PD-(L)1** | 933 (79.5) | 374 (76.6) | 1,307 (78.7) |
| **ANTI-CTLA-4** | 57 (4.9) | 42 (8.6) | 99 (6) |
| **ICI combination** | 183 (15.6) | 72 (14.8) | 255 (15.4) |
| **Most frequently found mutations (incidence >10% of all samples) N (%)** |  |  |  |
| **TP53** | 467 (39.8) | 271 (55.5) | 738 (44.4) |
| ***TERT*** | 256 (21.8) | 263 (53.9) | 519 (31.2) |
| ***KMT2D*** | 67 (5.7) | 169 (34.6) | 236 (14.2) |
| ***KRAS*** | 138 (11.8) | 88 (18) | 226 (13.6) |
| ***PIK3CA*** | 104 (8.9) | 96 (19.7) | 200 (12) |
| ***ARID1A*** | 59 (5.0) | 131 (26.8) | 190 (11.4) |
| ***NF1*** | 54 (4.6) | 129 (26.4) | 183 (11) |
| ***PTPRT*** | 48 (4.1) | 126 (25.8) | 174 (10.5) |

N = number; NSCLC = non-small cell lung cancer; MSI = microsatellite instability; ICI = immune checkpoint inhibitor; ANTI-PD-(L)1 = monoclonal antibody directed against programmed cell death protein-1 or monoclonal antibody directed against programmed cell death ligand 1; ANTI-CTLA-4 = monoclonal antibody directed against cytotoxic T lymphocyte-associated antigen; ICI COMBINATION = combined treatment regimen including one ANTI-PD-(L)1 and one ANTI-CTLA-4;

**Figure 1.**
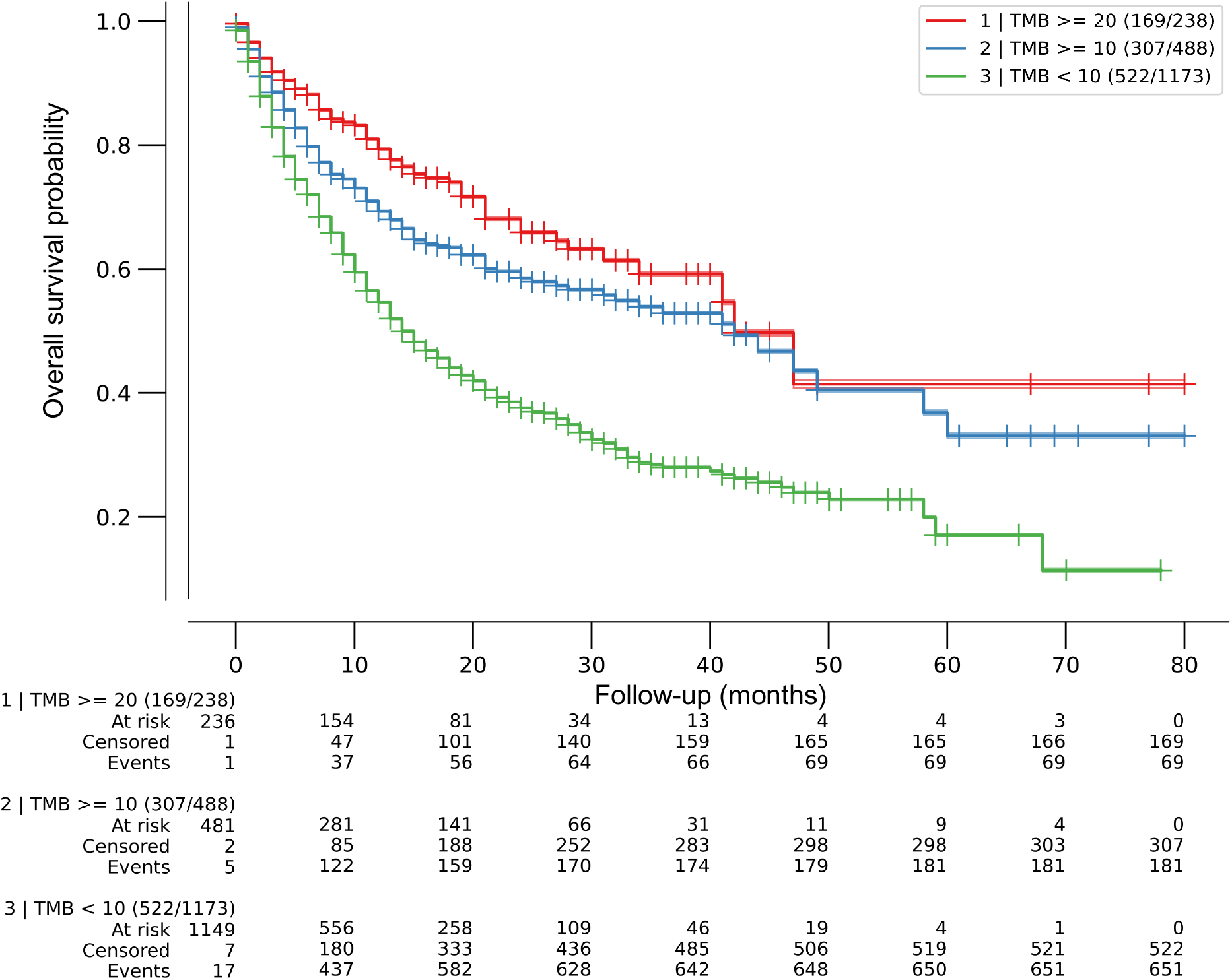
Effect of tumor mutational burden (TMB) on overall survival after ICI treatment. Kaplan–Meier (KM) curves for patients with tumors within each TMB predefined cutoff. Overall survival is from the first dose of ICI. Median OS was 42 months for both TMB ≥ 20mut/Mb and TMB ≥ 10 mut/Mb, and 15 months for TMB < 10 mut/Mb, multivariate log-rank P < 0.005. Cox regression HRs for death were 0.44 (95% CI 0.34 - 0.56) and 0.57 (95% CI 0.49 - 0.67) for TMB ≥ 20 mut/Mb and ≥ 10 mut/Mb, respectively (P < 0.005). No difference was observed in death risk between cohorts with TMB 10 mut/Mb or less and 1 mut/Mb or less (HR 0.96; 95% CI 0.74 – 1.24; P = 0.73).

### Single gene alterations and implications for survival

A total of 392 genes whose mutations were found in at least 5 patients were eligible for this analysis. Twenty-seven genes exhibited a statistically significant correlation with OS after ICI treatment when only tumors with TMB ≥ 10 mut/Mb were analyzed. Among them, 5 genes showed reduced OS on ICI (*STK11, KEAP1, CIC, E2F3*, and *TP53*), whereas 22 genes were associated with better OS (*NTRK3, TERT, NOTCH3, RNF43, TET1, PTPRD, NCOA3, TENT5C, ZFHX3, RIT1, CCNE1, PPM1D, GATA2, ALK, DNMT1, PTPRT, MET, EPHA7, BCL6, SMO, CDK6*, and *MED12)*, all P < 0.05. Individual OS for each gene is available in **Appendix Table A2**.

### Multivariate analysis of individual gene alterations in high TMB patients

Among the 27 genes exhibiting a correlation with OS (P < 0.05) at the univariate assessment, multivariate analysis confirmed a correlation between mutations in *STK11* (N=40, HR 1.84 (95% CI, 1.14 - 2.97) and *E2F3* (N=14, HR 3.17 (95% CI, 1.58 - 6.38]) and worse survival (P < 0.05). Mutations in *NTRK3* (N=57, HR 0.39 95% CI, 0.20 - 0.78), *PTPRD* (N=125, HR 0.67 95% CI, 0.45 - 0.99), *RNF43* (N=52, HR 0.42 95% CI, 0.2015 - 0.89), *TENT5C* (N=15, HR 0.14 95% CI, 0.02 - 0.98), *TET1* (N=55, HR 0.48 95% CI, 0.25 - 0.91), *and ZFHX3* (N= 91, HR 0.62 95% CI, 0.39 - 0.99) were associated with better OS. When evaluated concurrently with the mutational profile, histology did not play a relevant role in the ICI response, except for melanoma, which endowed patients with better OS (P < 0.05). TMB above the median TMB of the cohort (20mut/Mb) was also related to a better OS for all gene mutations (P < 0.05). In addition, MSI status, age, and gender did not have a consistent statistically significant effect on OS - **Figure 2**. KM curves for each gene related with survival after Cox multivariate analysis can be found in **Appendix Fig A3**.

**Figure 2.**
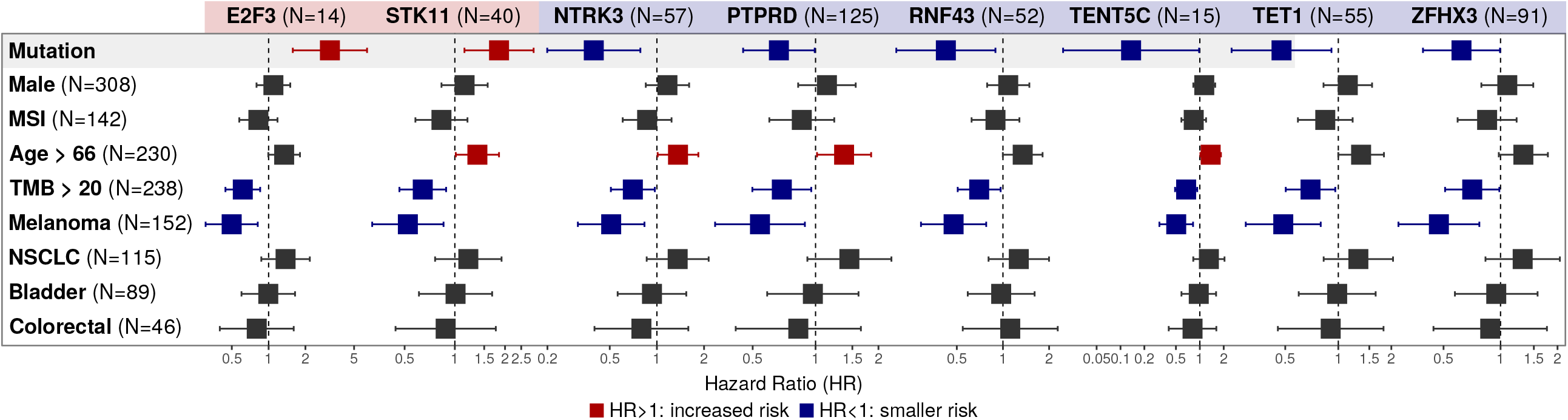
Multivariate analysis of individual gene alterations in high TMB patients. Forest plot for overall survival OS in multivariate analysis. Adjustment variables included TMB under or above the median of 20 mut/Mb, sex, median age, microsatellite instability (MSI) status, and tumor types (non-small cell lung cancer (NSCLC), melanoma, bladder cancer, and colorectal cancer). Mutations in *STK11* (N=40, HR 1.84 (95% CI, 1.14 - 2.97) and *E2F3* (N=14, HR 3.17 (95% CI, 1.58 - 6.38]) were related to worse survival (P < 0.05), while mutations in *NTRK* (N=57, HR 0.39 95% CI, 0.20 - 0.78), *PTPRD* (N=125, HR 0.67 95% CI, 0.45 - 0.99), *RNF43* (N=52, HR 0.42 95% CI, 0.2015 - 0.89), *TENT5C* (N=15, HR 0.14 95% CI, 0.02 - 0.98), *TET1* (N=55, HR 0.48 95% CI, 0.25 - 0.91), *and ZFHX3* (N= 91, HR 0.62 95% CI, 0.39 - 0.99) were associated with better OS. Melanoma histology and TMB above the median TMB of the cohort (20 mut/Mb) endowed patients with better OS (P < 0.05). MSI status, age, and gender did not have a consistent statistically significant effect on OS.

## DISCUSSION

Despite prior data demonstrating that higher TMB correlates with responses to ICIs, the use of TMB as a predictive biomarker for OS still has limitations^3^. In the original publication^1^, both the TMB evaluation as a continuous variable and the binary cutoff of the top 20% TMB within each histology with adjustment for cancer type, age, drug class of ICI, and year of ICI start reduced the chance of death across multiple cancer types (HR of 0.99 and 0.61, respectively, P□< 0.01). This data came across our findings, which validated and reinforced the predictive value of TMB ≥ 10 mut/Mb (HR of 0.57, P < 0.005) for response to ICI treatment. Also, a stricter TMB cutoff of ≥ 20 mut/Mb could refine patient selection (HR 0.44, P < 0.005). Conversely, visiting Marabelle *et al*.^2^, only a not-statistically significant difference in estimated 3-year OS was noted between the TMB-high/low groups (32% vs 22%), and more than half of the patients died regardless of TMB status at the 3-months landmark. In both cases, the use of additional clinical and molecular features could have provided better patient selection for ICI treatment.

While the Keynote-158 pivotal study that led to Pembrolizumab regulatory approval included small numbers of less common and often immune-refractory tumors (anal, biliary, cervical, endometrial, mesothelioma, neuroendocrine, salivary, thyroid, vulvar and small-cell lung cancers), the study cohort evaluated in our study reflects the real world epidemiology of ICIs use for more frequent tumors as NSCLC, melanoma, bladder cancer, and colorectal cancer.

Regarding the gene mutations, some of them were still less frequent, and the statistical positive association could not be translated to the clinical scenario. Examining examples separately, *CCND3* mutation occurred in 2 cases: one patient with melanoma (concurrent *KIT* and *BRAF* non-V600 mutations) and a second with NSCLC (concurrent *TP53 mutation*), and within two months of follow-up both patients died. Mutation in *TIMM8B* occurred in only one subject with colorectal cancer (TMB of 52.14 mut/Mb and concurrent *BRAF* V600E mutation), and the patient died within one month of PD-1 inhibitor monotherapy. Mutations in *CDKN2B* occurred in one case of cancer of unknown primary site (with concurrent *TP53* mutation), one patient with colorectal cancer, and a third case with melanoma. Within 8 months of follow-up, all patients were dead. Although the selection of mutations occurring in n > 5 patients for the univariate analysis was arbitrary, we aimed to increase data liability. Also, the complexity of antitumor immune responses reflexes in the absence of a universal biomarker to predict survival benefit from ICI^11^ and an integrative analysis of clinical and molecular variables may guide better patient selection for ICI treatment^12,13^.

Concerning MSI status, Goodman *et al*^14^. prior evaluated 148,803 tumor samples for TMB and MSI status. Overall, 18.3% of TMB-high tumors harbored MSI, which is significantly less than the findings of 29.1% in our cohort. Although, in both cases, microsatellite stable (MSS)/TMB-high amounts for a subgroup of cancers considerably larger than the MSI subset. Exploratory univariate analyses performed in our TMB-high cohort identified that patients with TMB-high/MSI tumors exhibit better OS outcomes when compared with TMB-high/MSS tumors (median OS 42 vs 19 months; P < 0,05). Cox HR for death was 0.77 (95% CI 0.60-0.98; P = 0.04) – **Appendix Fig A4 e A5**. Further, multivariate analysis to detect MSI influence on survival in TMB-high tumors was negative for all genes listed, which could hypothesize the presence of response prediction hierarchy greater for specific gene mutations than MSI. These findings were supported by the recently presented results of the CheckMate-848 study^15^, a phase 2 trial that tested Nivolumab with or without Ipilimumab for advanced or metastatic TMB-high solid tumors’ treatment. Exploratory analysis of the trial regarding the ORR among the TMB-high cohort showed that the responses were observed regardless of the MSI status. MSI was detected in 25 (2.5%) of MSI-evaluable tissue samples. ORR for these patients ranged from 33.3% to 55.6% with Nivolumab and Nivolumab plus Ipilimumab respectively. For MSS tumors, responses ranged from 26.9% to 29.1% with the same ICI regimens. Our multivariate analysis did not find any clinical feature, except for melanoma histology (19.3% of samples studied), that could interfere with survival outcomes after ICI. This melanoma enrichment could also justify a lack of correlation of survival and MSI status, as previous data shows the opposite^16^.

Notably, we identified two mutated genes related to poor survival: *STK11* and *E2F3*. Interestingly, there is stronger evidence supporting the STK11-related ICI resistance^17^. The STK11 mutation-induced downregulation of immune checkpoint regulating proteins like PD-L1 and T-cell chemokines favors a ‘‘cold’’ immunosuppressive tumor microenvironment (TME) and contributes to the exclusion of inflamed immune cells such as CD4+ T cells, CD8+ T cells, natural killer cells (NK), and Macrophage type 1 (M1), driving the tumor immune escape^18^. In accordance, the cell-cycle promoter *E2F3* is a well described tumoral poor prognosis factor and associated with a low immune signature score when amplified^19(p3)^.

Exploring gene mutations related to better OS, NTRK3 protein expression was previously positively associated with higher tumor immune and stromal scores, a great variety of immune lymphocytes, improved immune response, and ultimately with better survival. Accordingly, *NTRK3* may be a novel biomarker for ICI outcomes in selected tumor types^20(p3)^. The *PTPRD* gene encodes protein tyrosine phosphatase receptor type D (PTPRD) which is related to an increased mRNA expression of JAK1 and STAT1, subsequently attracting T cells through chemokines overexpression. Amidst NSCLC, *PTPRD* mutations endowed patients that received ICIs with better OS^21^. The effect of the mutation in *RNF43* gene in the TME was evaluated by Zhang *et al*.^22(p43)^ Their computed analysis of the tumor infiltrating immune cells identified an increment in CD8+ T cells, M1, NK, and total T cells compared with the wild type *RNF43* group. The *TENT5C gene* (which is interchangeable with *FAM46C*) was recently found to be a possible predictor of ICI efficacy as FAM46C expression was correlated with the abundance of CD4+ T cells, CD8+ T cells, and plasma B lymphocytes in the TME. Also, FAM46C expression was positively correlated with immune chemokines and immune chemokine receptors in most tumours^23^. For the *TET1* gene, previous studies found that tumor infiltrating lymphocytes, particularly the cytotoxic ones, were more abundant among the TET1-mutated tumors. Also, the neoantigen load was higher in this group, indicating that TET1 mutations were correlated with enhanced tumor immunogenicity^24(p1)^. Finally, for the *ZFHX3* gene, Zhang *et al*. hypothesized that *ZFHX3*-mutated tumors harbored increased expression of antigen-presentation-related molecules, stimulating immune-related ligands and receptors and chemokines. Also, mRNAs analysis of these tumors revealed a significantly increased immune checkpoint gene profiles^25^. All the mechanisms described above are potential explanations for our findings, and prospective literature data comprising this molecular selection for ICI use still lack.

Our study has limitations, including its retrospective nature based on clinical and molecular data available in a public online database and a bias toward hypermutated tumors (NSCLC, melanoma, and bladder cancer)^13^, which could explain the higher proportion of samples with TMB ≥ 10 mut/Mb (N 488, 29.4%). Second, some clinical information was not available, including objective responses to ICIs, preventing us from establishing associations. Third, we did not include a cohort of TMB-high patients not receiving ICIs; hence, a pure prognostic role of the genes described here cannot be ruled out and future studies can explore their predictive value. In addition, we used mutation in DNA repair genes to define MSI^6^, not being able to state that all these patients have microsatellite instability phenotypes.

## CONCLUSION

With this pan-cancer analysis, we demonstrated that not only high TMB, but also its combination with somatic mutational profile in some specific genes, can be a predictor of survival benefit from ICI treatment. Although prospective trials are still needed, combining information of TMB and mutation profiles in key cancer genes can be decisive in better qualify patients for ICI treatment.

## Data Availability

All data produced in the present study are available upon reasonable request to the authors.

## FIGURE LEGENDS

**Appendix Fig A3. Kaplan–Meier (KM) curves for each gene related with survival after Cox multivariate analysis**.

**Appendix Fig A4 and Appendix Fig A5. Effect of Microsatellite Instability (MSI) on overall survival after ICI treatment**.

Kaplan–Meier (KM) curves for TMB-high/MSI and with TMB-high/MSS tumors. Exploratory analyses performed in our TMB-high cohort identified that patients with TMB-high/MSI tumors exhibit better OS outcomes when compared with TMB-high/MSS tumors (median OS 42 vs 19 months; P < 0,05). Cox HR for death was 0.77 (95% CI 0.60-0.98; P = 0.04).

## AUTHOR CONTRIBUTIONS

Conception and design: Camila B. Xavier, Carlos Diego H. Lopes, Beatriz M. Awni, Eduardo F. Campos, and Denis L. Jardim.

Collection and assembly of data: Camila B. Xavier, Eduardo F. Campos, João Pedro B. Alves, Gabriela D. A. Guardia, Pedro A. F. Galante.

Manuscript writing: All authors.

Final approval of manuscript: All authors.

Accountable for all aspects of the work: All authors.

Provision of study materials or patients: All authors.

