## Appendix Fig A1. for "Interplay between Tumor Mutational Burden and Mutational Profile and its effect on overall survival: A Post Hoc Analysis of Metastatic Patients Treated with Immune Checkpoint Inhibitors"

**Supplementary Figure 1. Study design flowchart.**

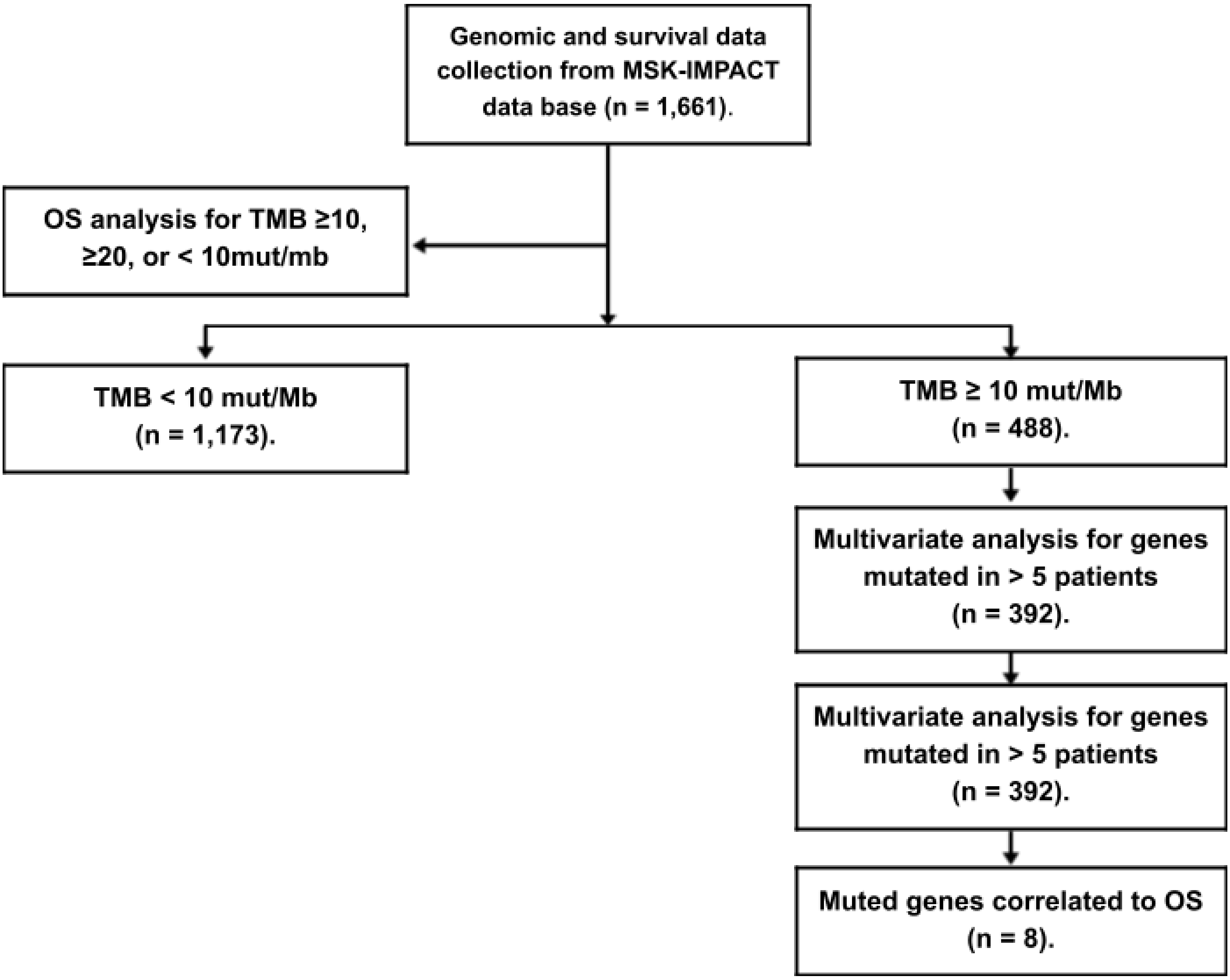


OS = overall survival; TMB = tumor mutational burden; mut/Mb = mutations per megabase; MSI = microsatellite instability
