## Appendix Table A2. for "Interplay between Tumor Mutational Burden and Mutational Profile and its effect on overall survival: A Post Hoc Analysis of Metastatic Patients Treated with Immune Checkpoint Inhibitors"

**Appendix Table A2. Individual survival data for genes at univariate analysis.**

| **Gene** | **Patients (N)** | **Median OS (months)** | **P value** |
| --- | --- | --- | --- |
| *STK11* | | | |
| *mut* | 40 | 7 | 0.00004 |
| *wt* | 448 | 44 |  |
| *KEAP1* | | | |
| *mut* | 52 | 13 | 0.002 |
| *wt* | 436 | 44 |  |
| *CIC* | | | |
| *mut* | 49 | 41 | 0.02 |
| *wt* | 439 | 42 |  |
| *E2F3* | | | |
| *mut* | 14 | 12 | 0.03 |
| *wt* | 474 | 44 |  |
| *TP53* | | | |
| *mut* | 271 | 28 | 0.03 |
| *wt* | 217 | 47 |  |
| *NTRK3* | | | |
| *mut* | 57 | NR | 0.0009 |
| *wt* | 431 | 36 |  |
| *TERT* | | | |
| *mut* | 263 | 49 | 0.002 |
| *wt* | 225 | 34 |  |
| *NOTCH3* | | | |
| *mut* | 77 | 47 | 0.003 |
| *wt* | 411 | 41 |  |
| *RNF43* | | | |
| *mut* | 52 | NR | 0.004 |
| *wt* | 436 | 36 |  |
| *TET1* | | | |
| *mut* | 55 | NR | 0.005 |
| *wt* | 433 | 36 |  |
| *PTPRD* | | | |
| *mut* | 125 | NR | 0.006 |
| *wt* | 363 | 41 |  |
| *NCOA3* | | | |
| *mut* | 28 | NR | 0.008 |
| *wt* | 460 | 41 |  |
| *TENT5C* | | | |
| *mut* | 15 | NR | 0.008 |
| *wt* | 473 | 41 |  |
| *ZFHX3* | | | |
| *mut* | 91 | NR | 0.01 |
| *wt* | 397 | 41 |  |
| *RIT1* | | | |
| *mut* | 11 | NR | 0.02 |
| *wt* | 477 | 42 |  |
| *CCNE1* | | | |
| *mut* | 9 | NR | 0.03 |
| *wt* | 479 | 42 |  |
| *PPM1D* | | | |
| *mut* | 21 | NR | 0.03 |
| *wt* | 467 | 42 |  |
| *GATA2* | | | |
| *mut* | 12 | NR | 0.03 |
| *wt* | 476 | 42 |  |
| *ALK* | | | |
| *mut* | 73 | 42 | 0.03 |
| *wt* | 415 | 36 |  |
| *DNMT1* | | | |
| *mut* | 39 | NR | 0.04 |
| *wt* | 449 | 41 |  |
| *PTPRT* | | | |
| *mut* | 126 | 44 | 0.04 |
| *wt* | 362 | 36 |  |
| *MET* | | | |
| *mut* | 38 | NR | 0.04 |
| *wt* | 450 | 41 |  |
| *EPHA7* | | | |
| *mut* | 76 | NR | 0.04 |
| *wt* | 412 | 41 |  |
| *BCL6* | | | |
| *mut* | 18 | NR | 0.04 |
| *wt* | 470 | 41 |  |
| *SMO* | | | |
| *mut* | 32 | NR | 0.04 |
| *wt* | 456 | 46 |  |
| *CDK6* | | | |
| *mut* | 8 | NR | 0.04 |
| *wt* | 480 | 42 |  |
| *MED12* | | | |
| *mut* | 38 | NR | 0.04 |
| *wt* | 450 | 42 |  |

N = number; OS = overall survival; NR = not reached
