## Supplementary figures and images for "Interplay between Tumor Mutational Burden and Mutational Profile and its effect on overall survival: A Post Hoc Analysis of Metastatic Patients Treated with Immune Checkpoint Inhibitors"

### Appendix Fig A3.

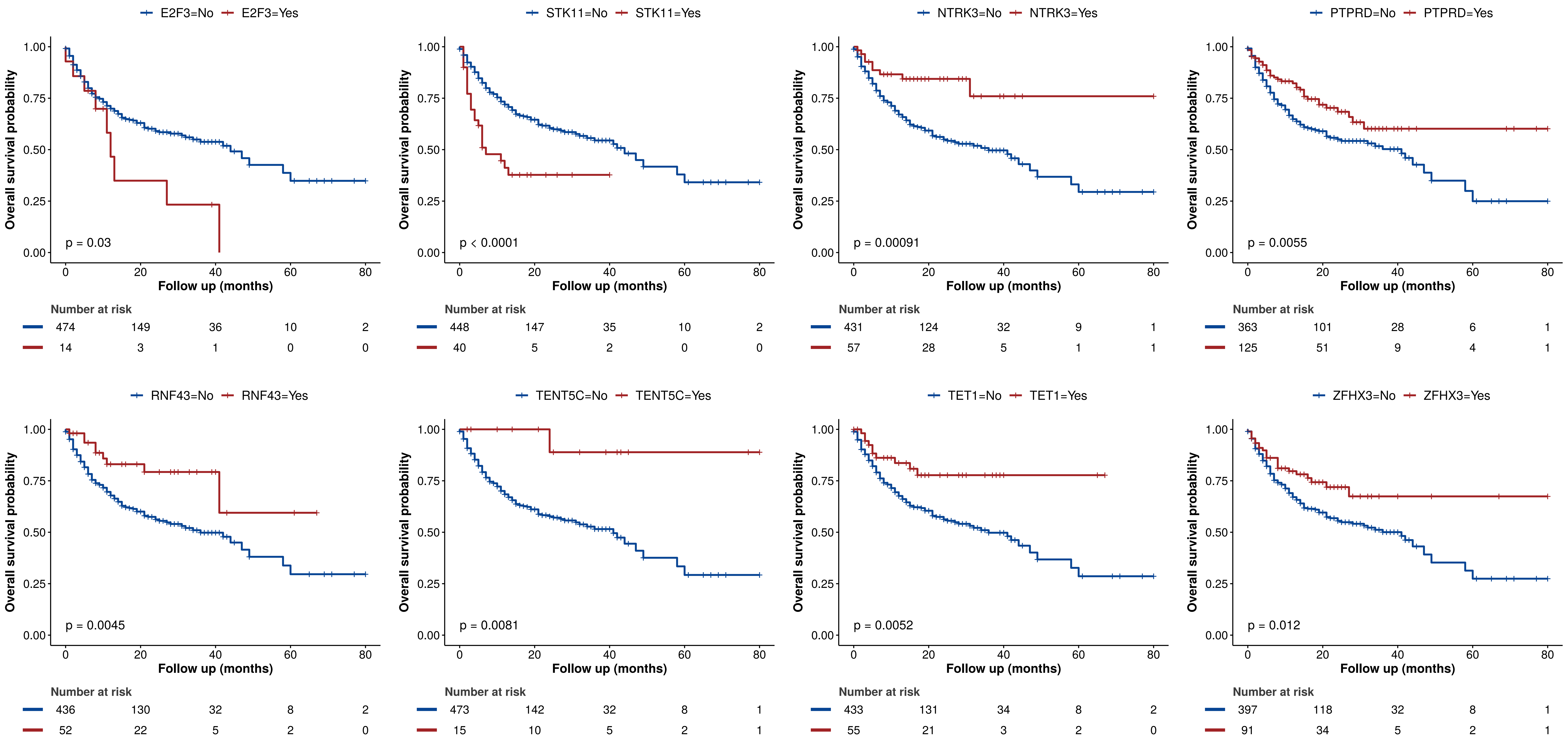

### Appendix Fig A4.

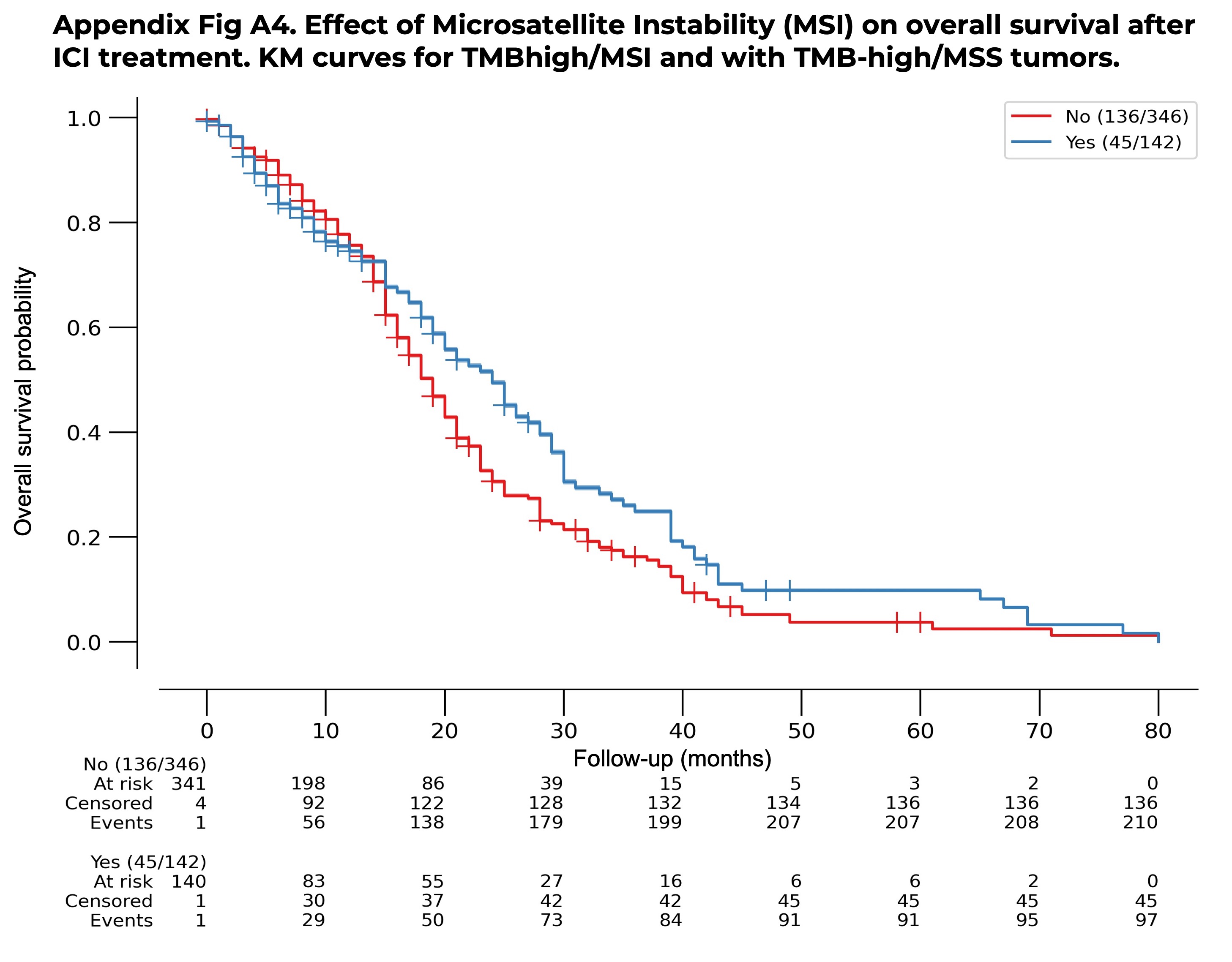

### Appendix Fig A5.

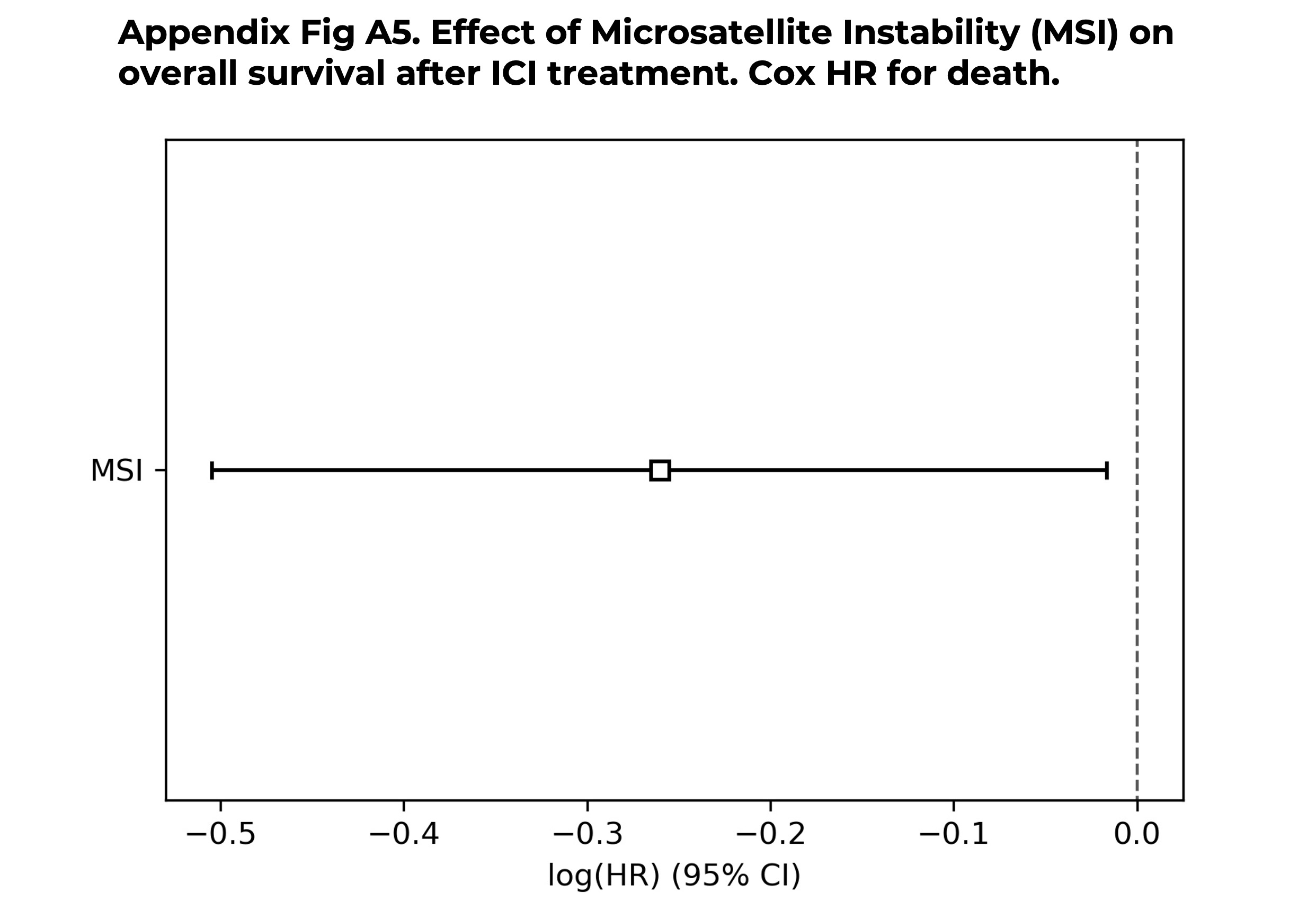
